# Data-driven identification of health conditions associated with incident Alzheimer’s disease dementia risk: a 15 years follow-up cohort from electronic health records in France and the United Kingdom

**DOI:** 10.1101/2021.06.07.21258454

**Authors:** Thomas Nedelec, Baptiste Couvy-Duchesne, Fleur Monnet, Timothy Daly, Manon Ansart, Laurène Gantzer, Béranger Lekens, Stéphane Epelbaum, Carole Dufouil, Stanley Durrleman

**Author notes:** Corresponding author: Carole Dufouil, PhD, Inserm u1219 “Bordeaux Population Health”, Bordeaux University, 146 rue Léo Saignat, 33076 Bordeaux. equal contributions (Carole Dufouil, Stanley Durrleman).

## Abstract

**Importance:** The identification of modifiable risk factors for Alzheimer’s disease (AD) is paramount for early prevention and the targeting of new interventions.

**Objective:** To assess the associations between health conditions diagnosed in primary care and the risk of incident AD over time.

**Design, Setting, and Participants:** Data for 20,214 AD patients from the United Kingdom and 19,458 AD patients from France were extracted from The Health Improvement Network (THIN) database. For each AD case, a control was randomly assigned after matching for sex and age at dementia diagnosis. We agnostically tested the associations between 123 different ICD10 diagnoses extracted from health records and AD dementia, by conditional logistic regression. We focused on two time periods: 2 to 10 years vs. 0 to 2 years before the diagnosis of AD, to separate risk factors from early symptoms/comorbidities.

**Exposures:** We considered all health conditions that had been recorded in more than 0.1% of visits per 1000 person-years in both cohorts, corresponding to 123 potential types of exposure.

**Main Outcomes and Measures:** Odds ratios (ORs) for the association of AD with the various health conditions were calculated after Bonferroni correction for multiple comparisons.

**Results:** Ten health conditions were significantly associated with high odds ratios for AD when diagnosed 2 to 10 years before AD, in the British and French samples: major depressive disorder (OR 95% confidence interval (UK):1.23-1.46)), anxiety (1.25-1.47), reaction to severe stress (1.24-1.59), hearing loss (1.11-1.28), constipation (1.22-1.41), spondylosis (1.14-1.39), abnormal weight loss (1.33-1.63), malaise and fatigue (1.14-1.32), memory loss (6.65-8.76) and syncope and collapse (1.1-1.37). Depression was the first comorbid condition associated with AD, appearing at least nine years before the first clinical diagnosis of AD, followed by, anxiety, constipation and abnormal weight loss.

**Conclusions and Relevance:** These results from two independent primary care databases provide new evidence on the temporality of risk factors and early signs of Alzheimer’s disease. These results could guide new dementia prevention strategies.

## Introduction

Alzheimer’s disease (AD) is thought to account for 60-70% of dementia cases worldwide, and is, thus, one of the principal health challenges of the 21^st^ century (WHO, 2020). Dementia affected 10.5 million Europeans in 2015^23^, and projections suggest that 13.5 million Europeans will be affected in 2030^23^. It is estimated that 4.5% of people over the age of 65 years currently have AD in Europe (6.4% for dementia in general)^23^. AD has major consequences at the individual and societal levels, as it requires a high degree of social care^21^.

The neurodegenerative diseases responsible for dementia (and AD in particular) are progressive and develop over decades before becoming disabling. Attempts to develop an effective disease-modifying treatment for patients with symptomatic AD have been unsuccessful. There has, therefore, been a shift towards early interventions, to maximise the therapeutic window through primary prevention measures acting on actionable risk factors, or secondary prevention measures to slow disease progression through early therapeutic interventions. The key to the implementation of such preventive measures is a data-driven understanding of the complex dynamics of the presymptomatic period at the point of care.

### Review of potential risk factors for dementia

The most recent Lancet Commission on Dementia, Prevention, Intervention and Care ^20^ added three new modifiable risk factors (excessive alcohol consumption^33^, head injury^36^ and air pollution^27^) to the initial list of nine factors ^21^ (low level of education, hypertension, hearing impairment, smoking, obesity, depression, physical inactivity, diabetes and infrequent social contact). The report concluded that modifying these 12 risk factors might prevent or delay dementia in up to 40% of cases (7% for low level of education and 8% for hearing loss). For Alzheimer’s dementia, another recent large meta-analysis of 243 observational prospective studies and 153 randomised controlled trials ^35^ identified 10 modifiable risk factors: diabetes, hyperhomocysteinaemia, poor BMI management, low level of education, hypertension in midlife, orthostatic hypotension, head trauma, poor cognitive activity, stress and depression. All the studies stressed the importance of a specific feature of neurodegenerative diseases, which include a prodromal phase lasting up to 20 years before clinical diagnosis ^10,34^. This makes it more difficult to separate possible early symptoms from causal risk factors when describing the natural course of the disease. It also highlights the urgent need for observational prospective studies with long follow-up periods, to investigate the long-term relationship between medical conditions and dementia onset ^34,35^.

### Investigating long-term risk factors observable in primary care: an agnostic approach

Epidemiological studies on the aetiology of AD have focused on hypothesis-driven searches for potential risk factors, in cohort studies on older people (mostly over the age of 60 years). By contrast, agnostic studies of risk factors test all possible associations exhaustively, in a similar fashion to genome-wide association studies for finding genotype-phenotype associations. They require large samples for the detection of statistically significant signals for associations. The constitution of such large samples is made possible by access to large electronic health record databases ^9,26^, facilitating complex longitudinal analyses and the testing of multiple hypotheses in well-powered studies.

We set up and analysed two large nested case-control studies drawn from two large independent primary care databases in the UK and France, extracted from The Health Improvement Network (THIN) database. Through this extensive study, we sought to confirm some well-known dementia risk factors and exhibit new candidates for consideration in future anti-dementia strategies.

## Methods

### Study design and participants

#### THIN database

We used The Health Improvement Network (THIN®) database^3^, a large standardised European database of fully anonymised and non-extrapolated electronic medical records collected from physicians by Cegedim and coded according to International Classification of Diseases, 10th Revision (ICD-10) codes. The THIN database complies with all current European data protection laws (General Data Protection Regulation (GDPR)) and adheres to the Observational Medical Outcomes Partnership (OMOP) model. The French data were collected from a pool of 2,000 general practitioners^14^ representative of the French population in terms of age, sex and living area. The UK data were collected from 400 general practices, representing around 6% of the UK population, selected for the THIN quality data recording scheme with Vision practice management software ^32^. Several reports have already shown that the electronically coded diagnoses in this database are representative of the UK general practice population in terms of demographics and type of consultation ^3,17^. For each patient, the diagnosis corresponding to each visit, the prescriptions made by the general practitioner (GP) and all other diagnoses associated with these prescriptions were available. Data for educational level and social status were not provided, to preserve anonymity.

#### Data extraction on the basis of ICD10 classification

For each patient, we had access to all the disease diagnoses made by the general practitioner, whether the main reason for the visit or associated with a prescription made by the GP. In the French cohort, we may not have had access to the full history of some patients, if they consulted several different general practitioners during the study. We defined past exposure to the health conditions considered, according to the 10th International Classification of Disease (ICD10). ICD10 codes were provided directly in the French database. For the UK database, ICD10 code was obtained by converting READ codes according to the correspondence provided by the UK Clinical Terminology Centre ^7^. We used the first three characters of the code, defining the category of the disease. In this exploratory approach, we assessed the association of AD with each of the health conditions defined by ICD10 codes recorded in more than 0.1% of visits per 1000 person-years in both countries (the exact list is provided in the supplementary material). *Study design*

We extracted two samples of patients diagnosed with AD from the French and UK electronic health records in the THIN database (dementia cases according to International Classification of Diseases, Ninth Revision, Clinical Modification [ICD-10-CM], codes = F00 and G30). In France, all patients with AD and at least two years of follow-up were extracted, whereas, in the UK, 35% of patients with AD and at least two years of follow-up (*n*=20654 from a total of 59912 AD patients with two years of follow-up in the database) were sampled with a Python random sampling routine. Age at AD onset was defined as age at the first record coded with an AD diagnosis. We considered data recorded from January 1996 to April 2020 in the UK and from September 1996 to February 2019 in France. For each country, a sample of control individuals, with no history of diagnosis of neurodegenerative diseases — Alzheimer’s disease (F00 and G30), Parkinson’s disease (G20), frontotemporal dementia (G31.0), dementia with Lewy bodies (G31.8), Huntington’s disease (G10), or multiple sclerosis (G35) — was drawn, matched with AD cases for age (± 1 year) at last record in the database and sex. Patients with less than two years of follow-up were excluded (eFigure 2). A summary table of the original data is provided in Table 1, and a flowchart (eFigure 1) is provided in the supplementary material. The study included 20,214 AD cases and matched controls for the UK cohort and 19,458 AD cases and matched controls for the French cohort (Table 1). Based on these initial cohorts, we designed several nested case-control analyses. For the investigation of risk factors and symptoms, we considered several time frames preceding AD diagnosis. We calculated odds ratios for three time periods between exposure to a particular health condition and dementia onset: 0 to 2 years, 2 to 10 years and 10 to 15 years. The index date was defined as the date of AD diagnosis for AD patients, and as the date of AD diagnosis of the matched AD patient for controls. For each analysis, we individually matched each AD patient with a control for sex, age at last visit (± 1 year) and years of follow-up, to ensure that cases and controls had been observed over the same lifespan, as explained in eFigure 2 provided in the supplementary material.

**Table 1:** Characteristics of patients with Alzheimer’s disease and controls. *The data shown are numbers (%) or medians (IQR)*.

|  | Full analytical cohorts |  | With >=2 years of retrospective data |  | With >=10 years of retrospective data |  | With >=15 years of retrospective data |  |
| --- | --- | --- | --- | --- | --- | --- | --- | --- |
|  | Alzheimer's disease(n=39672) | Controls (n=39672) | Alzheimer's disease (n=38996) | Controls (n=38996) | Alzheimer's disease (n=18234) | Controls (n=18234) | Alzheimer's disease (n=9180) | Controls (n=9180) |
| *UK* |  |  |  |  |  |  |  |  |
| Total | 20214 | 20214 | 19940 | 19940 | 13064 | 13064 | 7731 | 7731 |
| Sex: Women | 14107 (69%) | 14107 (69%) | 13924 (69%) | 13924 (69%) | 9217 (70%) | 9217 (70%) | 5567 (72%) | 5567 (72%) |
| Men | 6107 (31%) | 6107 (31%) | 6016 (31%) | 6016 (31%) | 3847 (30%) | 3847 (30%) | 2164 (28%) | 2164 (28%) |
| Age at index date | 81 (76-86) | 81 (76-86) | 81 (76-86) | 81 (76-86) | 82 (76-86) | 82 (76-86) | 82 (77-86) | 82 (77-86) |
| Person-years of data available before index date | 13 (9-18) | 10 (5-17) | 13 (9-18) | 11 (6-17) | 16 (13-20) | 16 (12-20) | 19 (17-22) | 19 (17-22) |
| Number of visits per year | 7 (4-11) | 7 (3-11) | 7 (4-11) | 7 (3-11) | 7 (4-11) | 7 (4-11) | 7 (4-11) | 7 (4-11) |
| *France* |  |  |  |  |  |  |  |  |
| Total | 19458 | 19458 | 19056 | 19056 | 5170 | 5170 | 1449 | 1449 |
| Sex: Women | 12442 (63%) | 12442 (63%) | 12186 (63%) | 12186 (63%) | 3291 (63%) | 3291 (63%) | 906 (62%) | 906 (62%) |
| Men | 7016 (37%) | 7016 (37%) | 6870 (37%) | 6870 (37%) | 1879 (37%) | 1879 (37%) | 543 (38%) | 543 (38%) |
| Age at index date | 80 (75-85) | 80 (75-85) | 80 (75-85) | 80 (75-85) | 82 (77-86) | 82 (77-86) | 83 (77-86) | 83 (77-86) |
| Person-years of data available before index date | 7 (4-10) | 5 (3-8) | 7 (4-10) | 5 (4-8) | 13 (11-15) | 12 (11-14) | 17 (16-18) | 16 (16-17) |
| Number of visits per year | 5 (3-8) | 4 (2-7) | 5 (3-8) | 4 (2-7) | 5 (3-8) | 5 (2-7) | 5 (3-7) | 5 (3-7) |

### Statistical analysis

We first used conditional logistic regression to analyse the association between each health condition and the diagnosis of AD. Bonferroni correction for multiple comparisons was applied. All analyses were conducted with the StatsModel and ScikitLearn libraries in Python. Values of *p* < 0.0004 in two-tailed tests were considered significant (*p*=0.05 with Bonferroni correction for 123 potential exposures). We estimated effect size globally and by time between health condition exposure and AD diagnosis, as follows: (0 - 2] years, (2 – 10] years, and (10-15] years. In this last category, no health condition was found to increase the risk of AD onset in both countries. In the appendix, we provide tables including all the odds ratios calculated in the conditional logistic regression analysis, a table comparing the prevalences of the tested exposures in controls from the two countries and figures showing the odds ratios of each significantly associated variable. We also show (alpha%-(1-alpha)%, alpha=0.05/ntests=0.05/123=0.0004) confidence intervals, which take into account the uncertainty due to multiple comparisons.

We determined the independent contributions of the different health conditions to the risk of AD, by performing a conditional logistic regression analysis including all the health conditions significantly and independently associated with an increase in AD risk. Assuming causality, we calculated a combined population attributable fraction associated with health conditions displaying significant association with AD.

We tried to identify the window of exposure of greatest importance for the various health conditions, by calculating the incidence of the health conditions concerned in the AD and matched control samples and following its change over time until the index date, for all conditions found to be significantly associated with AD in the (2-10 years] time window. We merged the French and the UK nested case-control samples for this analysis. We assumed that incidence follows a Poisson distribution and calculated the associated confidence intervals. We added five additional comorbid conditions (sleep disorders, hypotension, disorders of urinary system, hypothyroidism and fracture) associated with AD when diagnosed in the (0-2] years preceding AD diagnosis, which have also been reported as possible risk factors for dementia in previous studies. We plotted the change in incidence ratio for each comorbid condition with time to AD diagnosis. We smoothed the incidence ratio curve by fitting a quadratic polynomial, to facilitate interpretation of the plot. For each comorbid condition, we show the plot from the first time point at which the 95% confidence intervals of the corresponding incidence rates are separate.

## Results

Our initial full analytical cohorts included 20214 patients with a diagnosis of Alzheimer’s disease in the UK and 19458 AD patients in France; the study flowchart is provided in eFigure 1 in the supplementary materials.

Table 1 describes the age and sex distribution of patients with AD and their matched controls in the two countries. The median age at diagnosis was 81 years in the UK and 80 years in France. There were more women than men in both countries (69% in the UK and 63% in France). More follow-up data were available for the UK sample, with 13 person-years of data available before the index date in the full analytical cohorts (versus seven years in France). The median number of visits per year was also higher in the UK than in France (seven versus five). At two years before the index date, 19940 AD patients were included in the UK and 19056 in France; at 10 years, the corresponding numbers were 13064 and 5170; and at 15 years, they were 7731 and 1449. Age at diagnosis was not dependent on the number of years of data available before the index date in the UK, whereas, in France, AD patients with at least 10 years of retrospective data before diagnosis had a median age at the index date of 82 years (versus 80 years for the full analytical cohort). The mean number of visits per year was similar for the case and control groups in both countries. In the control groups, the health condition most frequently recorded in the databases of both countries was essential hypertension, with a prevalence of 53% in the UK and 65% in France, followed by cough (42%) and dorsalgia (41.6%) in the UK and by dorsalgia (37.9%) and hypercholesterolaemia (32%) in France (eTable1).

We agnostically tested 123 possible health conditions in the (0-2], (2-10] and (10-15] years before the diagnosis of Alzheimer’s disease (eTable 2). Only 10 of the 123 health conditions were significantly positively associated with the diagnosis of AD in both countries (Table 2 and Figure 1), after Bonferroni correction for the (2-10] year time window. None of these health conditions was significantly associated with the diagnosis of AD in the (10-15] years time frame. Spondylosis was the only health condition significantly associated with AD diagnosis in the (2-10] year period but not in the (0-2] period, this relationship being observed only in the UK. Cervical spondylosis accounted for 63% of the reported spondylosis cases in the UK and 60% in France. Another 12 health conditions were found to be significantly positively associated with AD in both countries in the (0-2] year time frame (eTable2 and eFigure 3). Five of the 10 health conditions identified as significantly associated with AD (anxiety, constipation, spondylosis, memory loss and abnormal weight loss) remained significant (*p* < 0.05) in the conditional logistic model containing all these health conditions (Table 3 and eTable 3 for the French analysis). They were also significant in the French model (eTable3). Assuming causality for depression, anxiety, spondylosis, constipation, hearing loss and reaction to severe stress, the potential attributable risk associated with these health conditions was 14% in France and 18% in the UK.

**Table 2:**
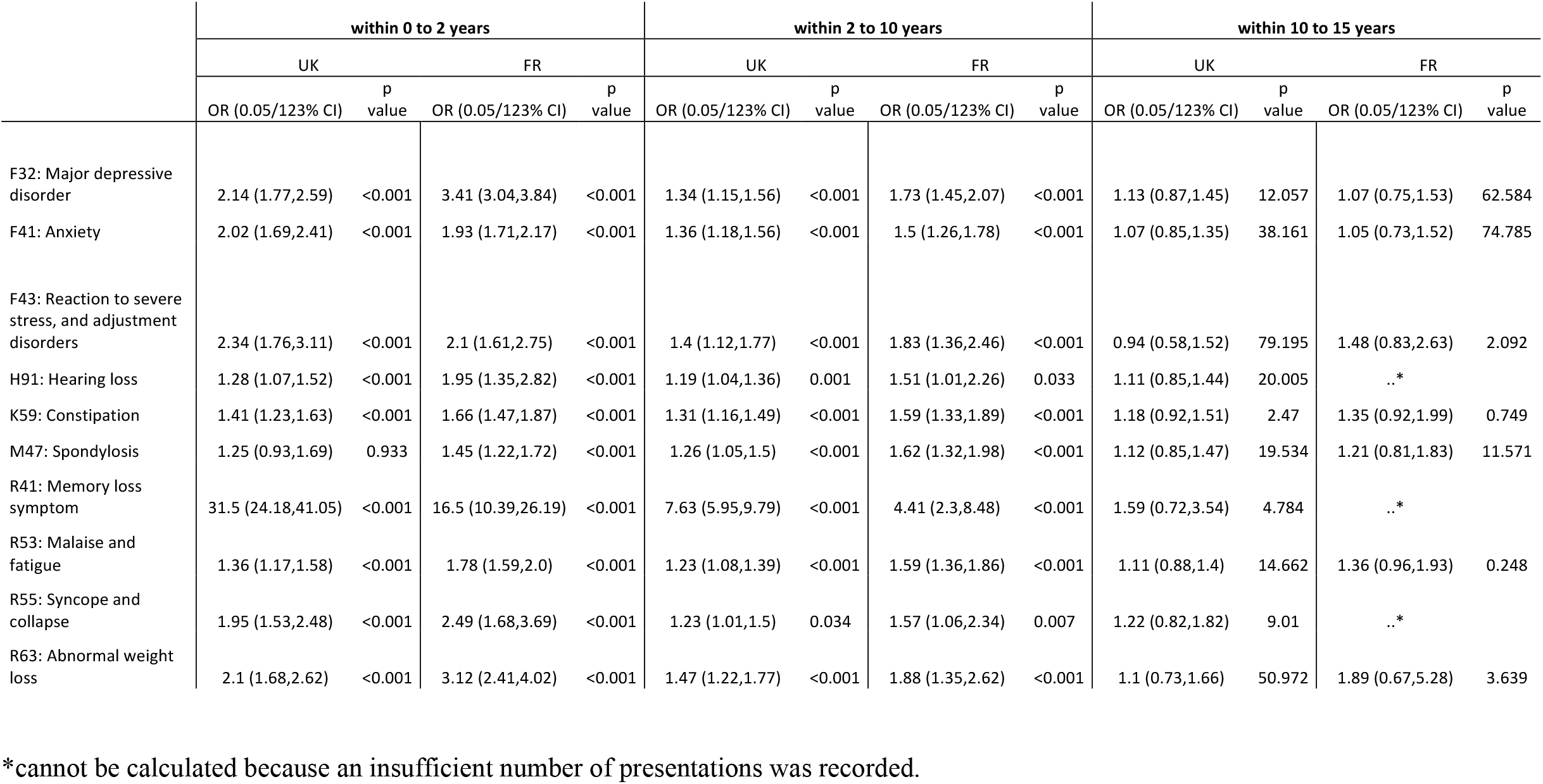
Odds ratios for all variables individually associated with a future diagnosis of AD in the (2-10] years before AD diagnosis. *We provide 95% confidence intervals corrected for multiple comparisons (i*.*e. (alpha%, 1-apha%) CI, with alpha = 0*.*05/123=0*.*0004) and associated p values corrected for multiple comparisons*.

|  | within 0 to 2 years |  |  |  | within 2 to 10 years |  |  |  | within 10 to 15 years |  |  |  |
| --- | --- | --- | --- | --- | --- | --- | --- | --- | --- | --- | --- | --- |
|  | UK |  | FR |  | UK |  | FR |  | UK |  | FR |  |
|  | OR (0.05/123% CI) | p value | OR (0.05/123% CI) | p value | OR (0.05/123% CI) | p value | OR (0.05/123% CI) | p value | OR (0.05/123% CI) | p value | OR (0.05/123% CI) | p value |
| F32: Major depressive disorder | 2.14 (1.77,2.59) | <0.001 | 3.41 (3.04,3.84) | <0.001 | 1.34 (1.15,1.56) | <0.001 | 1.73 (1.45,2.07) | <0.001 | 1.13 (0.87,1.45) | 12.057 | 1.07 (0.75,1.53) | 62.584 |
| F41: Anxiety | 2.02 (1.69,2.41) | <0.001 | 1.93 (1.71,2.17) | <0.001 | 1.36 (1.18,1.56) | <0.001 | 1.5 (1.26,1.78) | <0.001 | 1.07 (0.85,1.35) | 38.161 | 1.05 (0.73,1.52) | 74.785 |
| F43: Reaction to severe stress, and adjustment disorders | 2.34 (1.76,3.11) | <0.001 | 2.1 (1.61,2.75) | <0.001 | 1.4 (1.12,1.77) | <0.001 | 1.83 (1.36,2.46) | <0.001 | 0.94 (0.58,1.52) | 79.195 | 1.48 (0.83,2.63) | 2.092 |
| H91: Hearing loss | 1.28 (1.07,1.52) | <0.001 | 1.95 (1.35,2.82) | <0.001 | 1.19 (1.04,1.36) | 0.001 | 1.51 (1.01,2.26) | 0.033 | 1.11 (0.85,1.44) | 20.005 | ..* |  |
| K59: Constipation | 1.41 (1.23,1.63) | <0.001 | 1.66 (1.47,1.87) | <0.001 | 1.31 (1.16,1.49) | <0.001 | 1.59 (1.33,1.89) | <0.001 | 1.18 (0.92,1.51) | 2.47 | 1.35 (0.92,1.99) | 0.749 |
| M47: Spondylosis | 1.25 (0.93,1.69) | 0.933 | 1.45 (1.22,1.72) | <0.001 | 1.26 (1.05,1.5) | <0.001 | 1.62 (1.32,1.98) | <0.001 | 1.12 (0.85,1.47) | 19.534 | 1.21 (0.81,1.83) | 11.571 |
| R41: Memory loss symptom | 31.5 (24.18,41.05) | <0.001 | 16.5 (10.39,26.19) | <0.001 | 7.63 (5.95,9.79) | <0.001 | 4.41 (2.3,8.48) | <0.001 | 1.59 (0.72,3.54) | 4.784 | ..* |  |
| R53: Malaise and fatigue | 1.36 (1.17,1.58) | <0.001 | 1.78 (1.59,2.0) | <0.001 | 1.23 (1.08,1.39) | <0.001 | 1.59 (1.36,1.86) | <0.001 | 1.11 (0.88,1.4) | 14.662 | 1.36 (0.96,1.93) | 0.248 |
| R55: Syncope and collapse | 1.95 (1.53,2.48) | <0.001 | 2.49 (1.68,3.69) | <0.001 | 1.23 (1.01,1.5) | 0.034 | 1.57 (1.06,2.34) | 0.007 | 1.22 (0.82,1.82) | 9.01 | ..* |  |
| R63: Abnormal weight loss | 2.1 (1.68,2.62) | <0.001 | 3.12 (2.41,4.02) | <0.001 | 1.47 (1.22,1.77) | <0.001 | 1.88 (1.35,2.62) | <0.001 | 1.1 (0.73,1.66) | 50.972 | 1.89 (0.67,5.28) | 3.639 |

\*cannot be calculated because an insufficient number of presentations was recorded.

**Figure 1:**
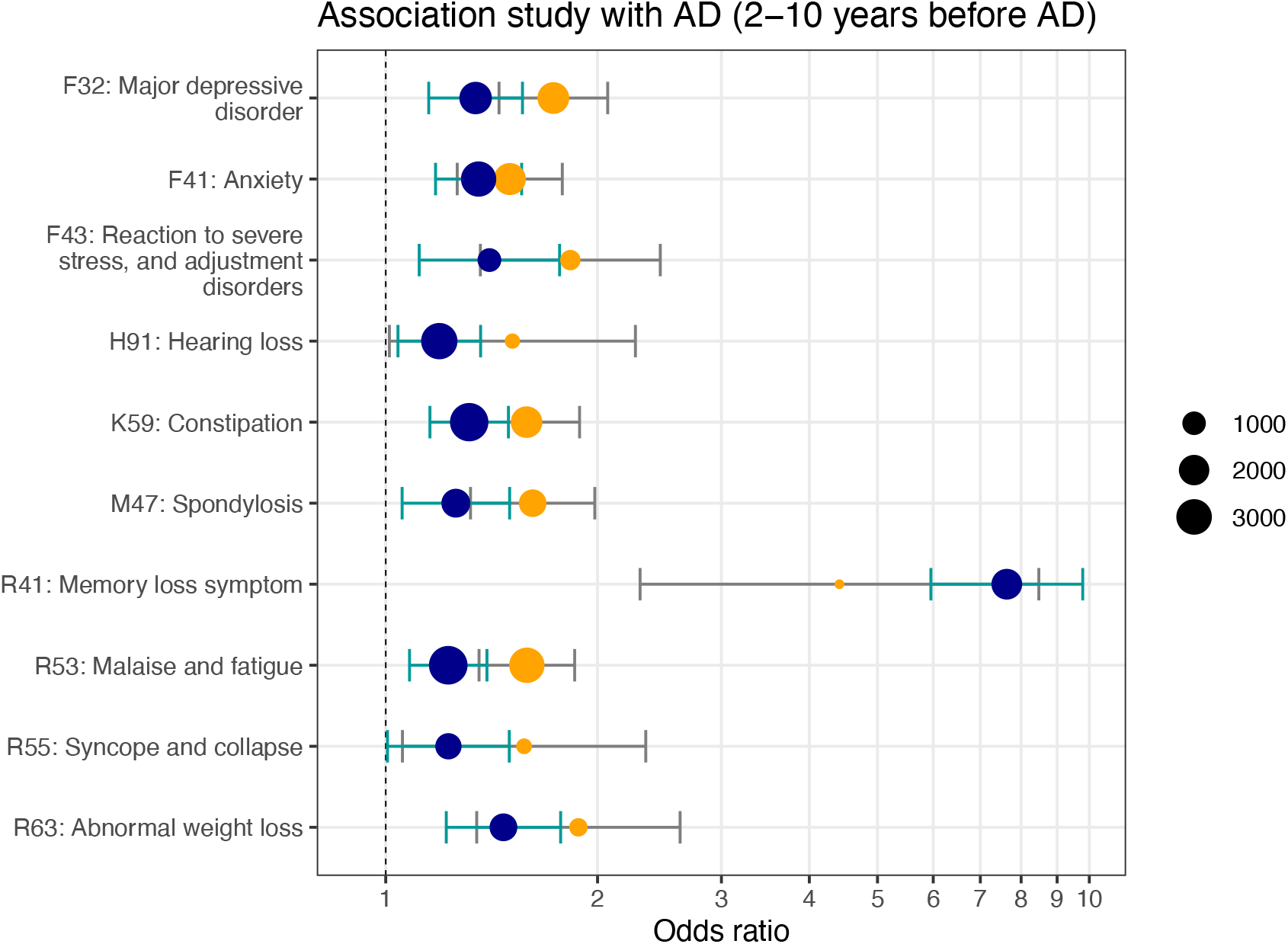
Health conditions associated with a future diagnosis of AD in the 2 to 10 years before diagnosis time period. *Only risk factors significant in both the French and UK cohorts are shown, with orange and blue dots, respectively. The size of the dot is proportional to the number of affected cases. Bars correspond to 95% CIs after correction for multiple comparisons*.

**Table 3:**
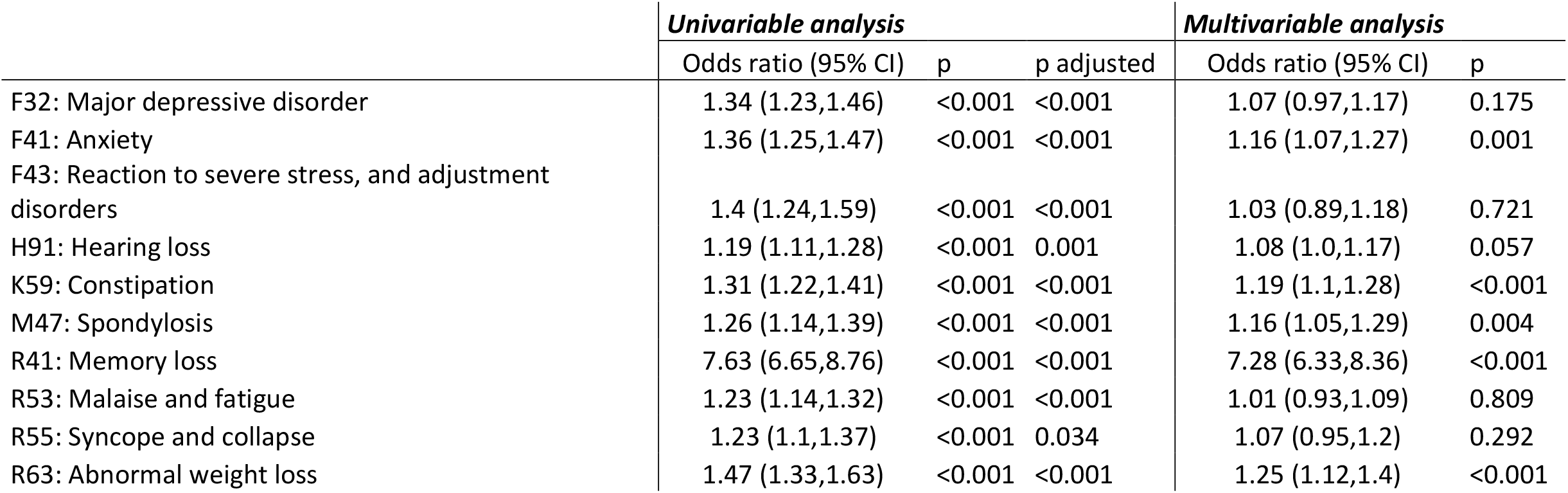
Odds ratio and 95% CIs for a conditional logistic model assessing the effect of all variables individually associated with a future diagnosis of AD in the UK.

|  | *Univariable analysis* |  |  | *Multivariable analysis* |  |
| --- | --- | --- | --- | --- | --- |
|  | Odds ratio (95% CI) | p | p adjusted | Odds ratio (95% CI) | p |
| F32: Major depressive disorder | 1.34 (1.23,1.46) | <0.001 | <0.001 | 1.07 (0.97,1.17) | 0.175 |
| F41: Anxiety | 1.36 (1.25,1.47) | <0.001 | <0.001 | 1.16 (1.07,1.27) | 0.001 |
| F43: Reaction to severe stress, and adjustment disorders | 1.4 (1.24,1.59) | <0.001 | <0.001 | 1.03 (0.89,1.18) | 0.721 |
| H91: Hearing loss | 1.19 (1.11,1.28) | <0.001 | 0.001 | 1.08 (1.0,1.17) | 0.057 |
| K59: Constipation | 1.31 (1.22,1.41) | <0.001 | <0.001 | 1.19 (1.1,1.28) | <0.001 |
| M47: Spondylosis | 1.26 (1.14,1.39) | <0.001 | <0.001 | 1.16 (1.05,1.29) | 0.004 |
| R41: Memory loss | 7.63 (6.65,8.76) | <0.001 | <0.001 | 7.28 (6.33,8.36) | <0.001 |
| R53: Malaise and fatigue | 1.23 (1.14,1.32) | <0.001 | <0.001 | 1.01 (0.93,1.09) | 0.809 |
| R55: Syncope and collapse | 1.23 (1.1,1.37) | <0.001 | 0.034 | 1.07 (0.95,1.2) | 0.292 |
| R63: Abnormal weight loss | 1.47 (1.33,1.63) | <0.001 | <0.001 | 1.25 (1.12,1.4) | <0.001 |

Finally, we show the change in incidence over time in the control and AD cohorts, for the health conditions significantly associated with AD risk (Figures 3 and 4). We found that depression and anxiety were the first health conditions to be associated with AD, at least nine years before the clinical diagnosis of AD, followed by constipation and abnormal weight loss seven years before the index date. These findings facilitate differentiation between risk factors associated with early stages of the disease and probable comorbid conditions occurring within a few years of diagnosis (e.g. sleep disorders, and urinary system disorders).

**Figure 2:**
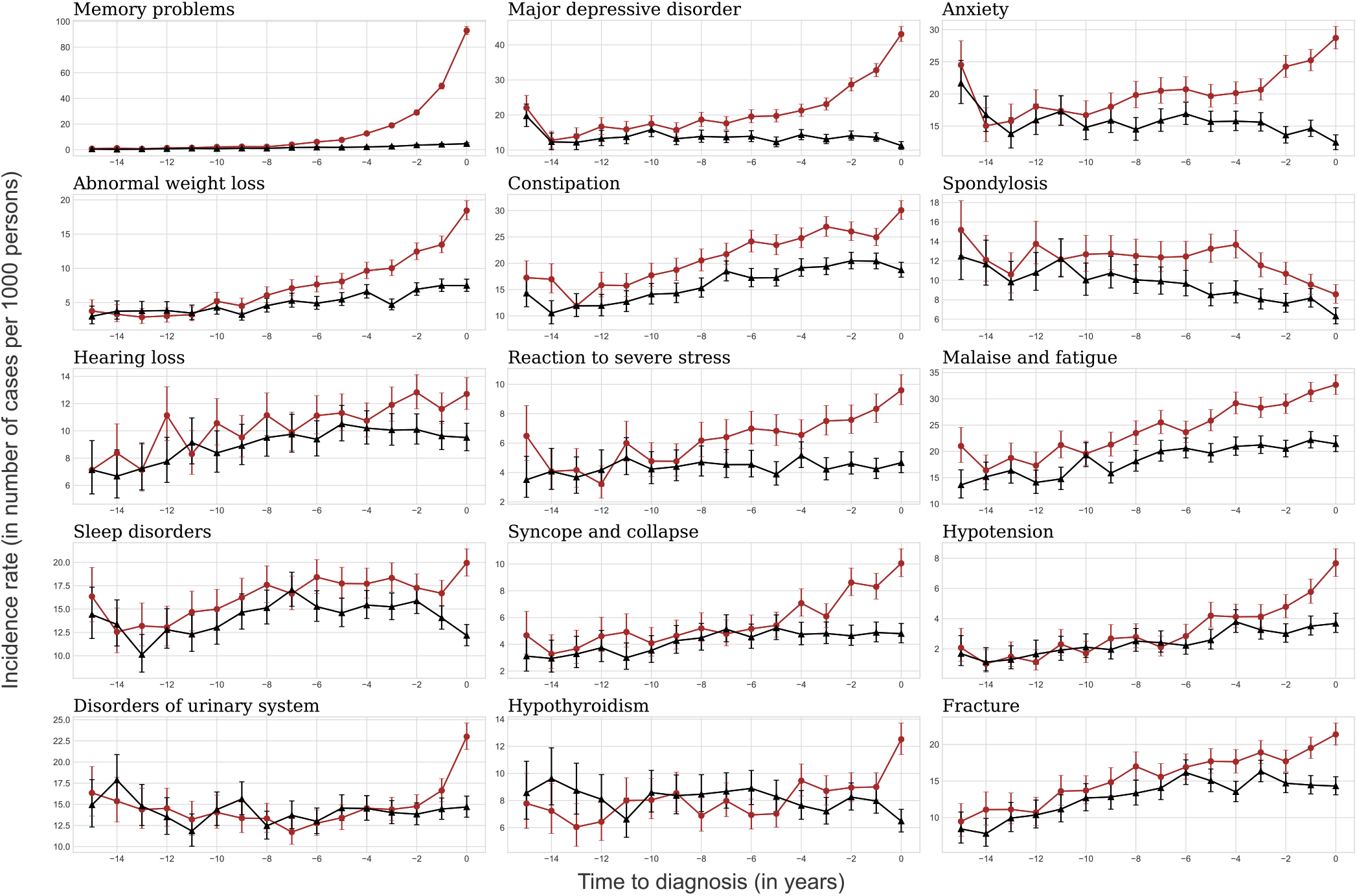
Incidence of comorbid conditions significantly associated with a subsequent AD diagnosis. The red curve corresponds to AD patients and the black curve corresponds to controls. Bars indicate the 95% CIs, based on the sample size of each group.

**Figure 3:**
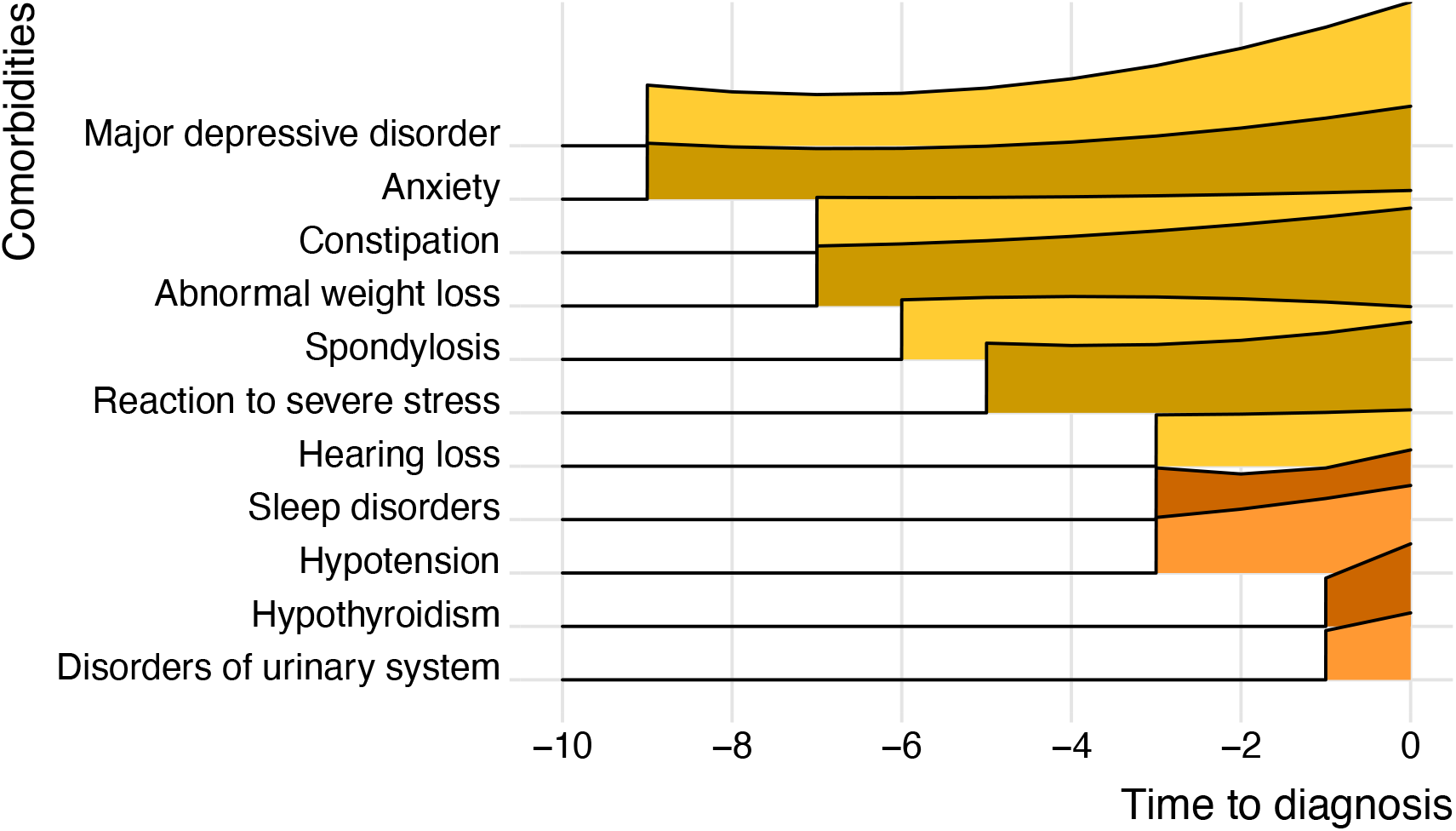
Change in incidence ratios over time for comorbid conditions significantly associated with AD. *Incidence ratios have been normalised and smoothed by quadratic regression*.

## Discussion

This large agnostic study of subjects from two European countries shows that several health conditions observed by general practitioners are significantly associated with the diagnosis of AD several years later. In these two studies, including a total of 39672 AD patients, 10 of the 123 health conditions considered were significantly associated with the subsequent diagnosis of AD in both countries and could be considered potential prediagnostic features. Depression and anxiety were among the risk factors for dementia identified by the recent Lancet Commission ^20^. Nevertheless, it remains a matter of debate whether these conditions are risk factors for dementia, early symptoms of dementia, or both^34^. The association between depression and subsequent AD diagnosis became significant at least nine years before the first clinical diagnosis of AD, consistent with the Whitehall II cohort study report^34^. Only anxiety was found to be associated with AD in the UK in a multivariable analysis of all the identified risk factors. This was the first multivariable analysis to consider all these health conditions at the same time.

Hearing loss^1,25^ has also been identified as a potential risk factor for dementia in multiple observational and prospective studies ^2,5,8,11,19^. A recent genetic study ^22^ found a correlation between genetically determined hearing impairment and AD. However, no causal link could be demonstrated between these traits, to confirm or infirm the current hypothesis that the simple management of hearing loss can decrease the risk of AD. We also observed a trend towards an association between AD and hearing loss, but it was not quite significant (*p*=0.057) after adjustment for other risk factors as possible cofounders (Table 3). We also confirmed associations between AD and abnormal weight loss^35^, which remained significant in the multivariable model. The causality of the link between abnormal weight loss and dementia remains to be demonstrated, and it is unclear whether this relationship is bidirectional.

The agnostic approach used here made it possible to identify other potential new risk factors for AD. Constipation has already been linked to depression^6^ and has been identified as a symptom in some neurodegenerative diseases, such as Lewy body dementia and Parkinson’s disease^32^. To our knowledge, this is the first time that constipation has been shown to be associated with a risk of AD. This association was detected seven years before the first clinical diagnosis of AD, and remained significant until the time of AD diagnosis and after controlling for depression and anxiety. We also found a strong association between spondylosis and AD dementia risk (OR=1.26 in the UK and OR=1.45 in France), which remained significant after correcting for other identified risk factors. One possible reason for this association may be a decline in physical activity^20^ due to spondylosis. A higher prevalence of dementia and AD was reported in a retrospective longitudinal study of patients with ankylosing spondylitis^16^. Nevertheless, we found an association with spondylosis (ICD10 code M47) but not with ankylosing spondylitis (ICD10 code M49). This result suggests that the association may not be due to the inflammatory nature of the spondylitis. We noted that the spondylosis was cervical in most of our cases. It may therefore be worthwhile investigating whether cervical spondylosis affects the flow of blood^29^ or cerebrospinal fluid^30^ to the brain, thereby favouring or accelerating neurodegeneration. The associations of AD with syncope and collapse and hypotension were also documented in a recent study reporting an association between midlife orthostatic hypotension and future dementia^28^.

We found associations present only in the last two years before diagnosis. The lateness of these associations suggests that these conditions are probably prodromal symptoms of the disease rather than risk factors. For instance, a recent Mendelian randomised study ^12^ showed that AD was more likely to be the cause of sleep disorders rather than a consequence of these disorders. The association with “hypothyroidism” may, therefore, reflect active investigations in patients with cognitive impairment, as the evaluation of thyroid hormone levels is recommended for this indication by the World Health Organisation.

We also highlight risk factors not replicated in the analysis. Hypertension has repeatedly been presented as a potential modifiable risk factor for AD^24^, whereas our data suggest a protective effect in the UK study and no association in the French study (FR: 1.01 (0.87-1.18), UK: 0.82 (0.75-0.90)). As in other observational studies ^4,13^, this association may result from a channelling effect, in which demented patients with hypertension or other cardiovascular comorbidities are more likely to be diagnosed with vascular dementia than AD.

Other previously reported associations not replicated in this study included associations with herpes virus infections, alcohol use disorders^33^, diabetic disorders and obesity. We cannot rule out the possibility that these discrepancies may be due to the control cohorts consisting of patients regularly consulting a general practitioner for these chronic illnesses.

### Strengths and limitations

Epidemiological approaches, such as Lancet Commissions ^20,21^, tend to focus on all-cause dementia, whereas other approaches in dementia research tend to argue for increasingly precise biological definitions of AD for the testing of hypothesis-driven specific pharmaceutical strategies ^15^. The early detection and labelling of biomarker-positive patients before the onset of symptoms has been criticised, due to the uncertainty of progression to dementia and the absence of proven risk-reducing interventions ^18,31^. This study has the advantage of bridging the gap between these different approaches, through a large-scale, exploratory, data-driven approach, considering all the main health conditions observable at the point of care.

Beyond its data-driven precision, another notable strength of this study is its inclusion of sufficient numbers of patients from two countries, making it possible to replicate the analysis, with consistent results obtained for the two samples, ensuring a high degree of generalisability. One question raised by this study is whether the variables identified are genuine risk factors. This could be tested in Mendelian randomisation studies on databases including genetic data^12^ or in new prospective research cohorts.

The study also has several limitations. We did not have access to other variables, such as education level, socio-economic status or genetic information. In addition, the recurrence of some diagnoses over time was not taken into account, as we considered only the first presentation of each health condition. Future studies could use the number of visits per diagnosis as a proxy for disease severity. The matched control samples were probably not entirely representative of the dementia-free general population, as they probably included a higher proportion of people with chronic illnesses or in poorer health generally. The odd ratios reported may therefore be an underestimate, as the real dementia-free general population includes a higher proportion of healthy ageing individuals than our control sample. The diagnosis of AD by a general practitioner rather than a trained neurologist may also have added to the background noise in the results, although this effect was probably counterbalanced by the large number of patients considered in the study. Diagnosis by a GP probably also resulted in a later diagnosis than in prospective cohorts in which each patient attends an annual visit with a neurologist. This is the main reason for which we considered comorbid conditions associated with AD at least the two years before the diagnosis of AD. We believe that the performance of a large number of studies of this type on large databases and their replication by independent research teams will lead to the emergence of a consensus for confirmation through future prospective cohorts.

### Conclusion

This study provides a modern extensive data-driven survey of potential risk factors and early symptomatic expression for Alzheimer’s disease in primary care, and constitutes a first attempt to determine when these comorbid conditions appear during the course of the disease. This study is innovative in its agnostic approach, leading not only to the confirmation of known risk factors for AD, but also to the identification of new risk factors. We confirmed several associations identified in previous studies, with hearing loss, depression and anxiety, for example, and identified other associations, with spondylosis, constipation and weight loss. Our findings are robust, as the associations were explored in two independent databases from France and the UK, ensuring the generalisability of the results. Finally, our findings make it possible to model the possible trajectories of risk factors in the period preceding the diagnosis of AD, providing new insight into possible windows for prevention.

## Data Availability

The data used in the preparation of the article are available from the Cegedim company upon reasonable request.

## Acknowledgement

This work was funded in part by the Agence Nationale de la Recherche, as part of the “Investissements d’avenir” program, under grants ANR-10-IAIHU-06 (IHU-ICM) and ANR-19-P3IA-0001 (PRAIRIE 3IA Institute). BCD is supported by a CJ Martin Fellowship (NHMRC, APP1161356).

## Contributors

TN, BCD, SE, CD and SD conceived and designed the study. FM, MA, LG, BL extracted the data. FM, MA and TN ensured data quality and TN, BCD, SE, CD and SD analysed the data. TN and BCD generated the figures. TN, BCD, SE, FM, MA, LG, BL, CD and SD interpreted the results and contributed to the writing of the final version of the manuscript. All authors agreed with the results and conclusions and approved the final draft.

## Ethics committee approval

The French database obtained approval for data collection from the French National Data Protection Authority (CNIL) in 2002. For the UK database, ethical approval was granted by the NHS South-East Multicentre Research Ethics Committee in 2003 (ref: 03/01/073) for establishment of the THIN database, and it was updated in 2011. The study was a retrospective analysis of secondary anonymised patient data only.

## Patient and Public involvement

We plan to disseminate the results of our research to the public and relevant patient community through the current publication and its related press release as well as presentations at scientific conferences.

The lead authors (CD,SD) affirm that this manuscript is an honest, accurate, and transparent account of the study being reported; that no important aspects of the study have been omitted; and that any discrepancies from the study as planned (and, if relevant, registered) have been explained.

